# Monitoring carbon dioxide to quantify the risk of indoor airborne transmission of COVID-19

**DOI:** 10.1101/2021.04.04.21254903

**Authors:** Martin Z. Bazant, Ousmane Kodio, Alexander E. Cohen, Kasim Khan, Zongyu Gu, John W. M. Bush

## Abstract

A new guideline for mitigating indoor airborne transmission of COVID-19 prescribes a limit on the time spent in a shared space with an infected individual (Bazant and Bush, 2021). Here, we rephrase this safety guideline in terms of occupancy time and mean exhaled carbon dioxide concentration in an indoor space, thereby enabling the use of CO_2_ monitors in the risk assessment of airborne transmission of respiratory diseases. While CO_2_ concentration is related to airborne pathogen concentration (Rudnick and Milton, 2003), the guideline developed here accounts for the different physical processes affecting their evolution, such as enhanced pathogen production from vocal activity and pathogen removal via face-mask use, filtration, sedimentation and deactivation. Critically, transmission risk depends on the total infectious dose, so necessarily depends on both the pathogen concentration and exposure time. The transmission risk is also modulated by the fractions of susceptible, infected and immune persons within a population, which evolve as the pandemic runs its course. A mathematical model is developed that enables a prediction of airborne transmission risk from real-time CO_2_ measurements. Illustrative examples of implementing our guideline are presented using data from CO_2_ monitoring in university classrooms and office spaces.

Impact Statement
There is mounting scientific evidence that COVID-19 is primarily transmitted through indoor airborne transmission, as arises when a susceptible person inhales virus-laden aerosol droplets exhaled by an infectious person. A safety guideline to limit indoor airborne transmission (Bazant and Bush, 2021) has recently been derived that complements the public health guidelines on surface cleaning and social distancing. We here recast this safety guideline in terms of total inhaled carbon dioxide, as can be readily monitored in most indoor spaces. Our approach paves the way for optimizing air handling systems by balancing health and financial concerns, informs policy for safely re-opening schools and businesses as the pandemic runs its course, and may be applied quite generally in the mitigation of airborne respiratory illnesses, including influenza.

## 1. Introduction

Coronavirus disease 2019 (COVID-19) has caused a devastating global pandemic since it was first identified in Wuhan, China in December 2019 (Chen et al., 2020; Li et al., 2020). For over a year, public health guidance has focused on disinfecting surfaces in order to limit transmission through fomites (Van Doremalen et al., 2020) and maintaining social distance in order to limit transmission via large drops generated by coughs and sneezes (Bourouiba et al., 2014). The efficacy of these measures has been increasingly called into question, however, since there is scant evidence for fomite transmission (Lewis, 2021) and large-drop transmission is effectively eliminated by masks (Moghadas et al., 2020).

There is now overwhelming evidence that the pathogen responsible for COVID-19, severe-acute-respiratory-syndrome coronavirus 2 (SARS-CoV-2), is transmitted primarily through exhaled aerosol droplets suspended in indoor air (Prather et al., 2020; Morawska and Milton, 2020; Morawska and Cao, 2020; Jayaweera et al., 2020; Zhang et al., 2020b; Bazant and Bush, 2021). Notably, airborne transmission provides the only rational explanation for the so-called “super-spreading events”, which have now been well chronicled and all took place indoors (Miller et al., 2020; Moriarty, 2020; Hamner, 2020; Shen et al., 2020; Nishiura et al., 2020; Kwon et al., 2020; Hwang et al., 2020). The dominance of indoor airborne transmission is further supported by the fact that face-mask directives have been more effective in limiting the spread of COVID-19 than either social distancing directives or lockdowns (Zhang et al., 2020b; Stutt et al., 2020). Indeed, a recent analysis of spreading data from Massachusetts public schools where masking was strictly enforced found no statistically significant effect of social distance restrictions that ranged from 3 feet to 6 feet (van den Berg et al., 2021). Finally, the detection of infectious SARS-CoV-2 virions suspended in hospital room air as far as 18 feet from an infected patient provides direct evidence for the viability of airborne transmission of COVID-19 (Lednicky et al., 2020; Santarpia et al., 2020).

With a view to informing public health policy, we proceed by developing a quantitative approach to mitigating the indoor airborne transmission of COVID-19, an approach that might be similarly applied to other airborne respiratory diseases. The canonical theoretical framework of Wells (1955) and Riley et al. (1978) describes airborne transmission in an indoor space that is well-mixed by ambient air flows, so that infectious aerosols are uniformly dispersed throughout the space (Gammaitoni and Nucci, 1997; Beggs et al., 2003; Nicas et al., 2005; Noakes et al., 2006; Stilianakis and Drossinos, 2010). While exceptions to the well-mixed room are known to arise (Bhagat et al., 2020), supporting evidence for the well-mixed approximation may be found in both theoretical arguments (Bazant and Bush, 2021) and computer simulations of natural and forced convection (Foster and Kinzel, 2021). The Wells-Riley model and its extensions have been applied to a number of super-spreading events and used to assess the risk of COVID-19 transmission in a variety of indoor settings (Miller et al., 2020; Buonanno et al., 2020b,a; Prentiss et al., 2020; Evans, 2020).

A safety guideline for mitigating indoor airborne transmission of COVID-19 has recently been derived that indicates an upper bound on the cumulative exposure time, that is, the product of the number of occupants and the exposure time (Bazant and Bush, 2021). This bound may be simply expressed in terms of the relevant variables, including the room dimensions, ventilation, air filtration, mask efficiency and respiratory activity. The guideline has been calibrated for COVID-19 using epidemiological data from the best characterized super-spreading events and incorporates the measured dependence of expiratory droplet-size distributions on respiratory and vocal activity (Morawska et al., 2009; Asadi et al., 2019, 2020a). An online app has facilitated its widespread use during the pandemic (Khan et al., 2020). The authors also considered the additional risk of turbulent respiratory plumes and jets (Abkarian et al., 2020a,b), as need be considered when masks are not worn. The accuracy of the guideline is necessarily limited by uncertainties in a number of model parameters, which will presumably be reduced as more data is analyzed from indoor spreading events.

Carbon dioxide measurements have been used for decades to quantify airflow and zonal mixing in buildings and so guide the design of heating, ventilation, and air-conditioning (HVAC) systems (Fisk and De Almeida, 1998; Seppänen et al., 1999). Such measurements thus represent a natural source of data for assessing indoor air quality, especially as they rely only on relatively inexpensive, widely available CO_2_ sensors. Quite generally, high carbon dioxide levels in indoor settings are known to be associated with poor health (Salisbury, 1986; Hung and Derossis, 1989; Seppänen et al., 1999). Statistically significant correlations between CO_2_ levels and illness-related absenteeism in both the work place (Milton et al., 2000) and classrooms (Mendell et al., 2013; Shendell et al., 2004) have been widely reported (Li et al., 2007). Direct correlations between CO_2_ levels and concentration of airborne bacteria have been found in schools (Liu et al., 2000). Correlations between outdoor air exchange rates and respiratory infections in dorm rooms have also been reported (Sun et al., 2011; Bueno de Mesquita et al., 2020). Despite the overwhelming evidence of such correlations and the numerous economic analyses that underscore their negative societal impacts (Milton et al., 2000; Fisk, 2000), using CO_2_ monitors to make quantitative assessments of the risk of indoor disease transmission is a relatively recent notion (Li et al., 2007).

Rudnick and Milton (2003) first proposed the use of Wells-Riley models, in conjunction with measurements of CO_2_ concentration, to assess airborne transmission risk indoors. Their model treats CO_2_ concentration as a proxy for infectious aerosols: the two were assumed to be produced proportionally by the exhalation of an infected individual and removed at the same rate by ventilation. The current pandemic has generated considerable interest in using CO_2_ monitoring as a tool for risk management of COVID-19 (Bhagat et al., 2020; Hartmann and Kriegel, 2020). The Rudnick-Milton model has recently been extended by Peng and Jimenez (2021) through consideration of the different removal rates of CO_2_ and airborne pathogen. They conclude by predicting safe CO_2_ levels for COVID-19 transmission in various indoor spaces, which vary by up to two orders of magnitude.

We here develop a safety guideline for limiting indoor airborne transmission of COVID-19 by expressing the safety guideline of Bazant and Bush (2021) in terms of CO_2_ concentration. Doing so makes clear that one must limit not only the CO_2_ concentration, but also the occupancy time. Our model accounts for the effects of pathogen filtration, sedimentation and deactivation in addition to the variable aerosol production rates associated with different respiratory and vocal activities, all of which alter the relative concentrations of airborne pathogen and CO_2_. Our guideline thus quantifies the extent to which safety limits may be extended by mitigation strategies such as mask directives, air filtration and the imposition of ‘quiet spaces’.

In §2, we rephrase the indoor safety guideline of Bazant and Bush (2021) in terms of the room’s carbon dioxide concentration. In §3, we present theoretical descriptions of the evolution of CO_2_ concentration and infectious aerosol concentration in an indoor space, and highlight the different physical processes influencing the two. We then model the disease transmission dynamics, which allows for the risk of indoor airborne transmission to be assessed from CO_2_ measurements taken in real time. In §4, we apply our model to a pair of data sets tracking the evolution of CO_2_ concentration in specific office and classroom settings. These examples illustrate how CO_2_ monitoring, when coupled with our safety guideline, provides a means of assessing and mitigating the risk of indoor airborne transmission of respiratory pathogens.

## 2. Safety Guideline for the Time-Averaged Carbon Dioxide Concentration

### 2.1. Occupancy-based safety guideline

We begin by recalling the safety guideline Bazant and Bush (2021) for limiting indoor airborne disease transmission in a well-mixed space. The guideline would impose an upper bound on the cumulative exposure time:

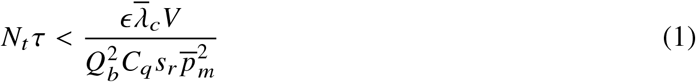

where *N*_*t*_ is the number of possible transmissions (pairs of infected and susceptible persons) and *τ* is the time in the presence of the infected person(s). The reader is referred to Table 1 for a glossary of symbols and their characteristic values. *Q*_*b*_ is the mean breathing flow rate and *V* the room volume. The risk tolerance *ϵ* < 1 is the prescribed bound on the probability of at least one transmission, as should be chosen judiciously according to the vulnerability of the population (Garg, 2020); for example, Bazant and Bush (2021) suggested *ϵ* = 10% for children and 1% for the elderly.

**Table 1.** Glossary of symbols arising in our theory, their units and characteristics values.

| Symbol | Meaning | Typical Values |
| --- | --- | --- |
| **Engineering Parameters** |  |  |
| $N$ | Number of persons, room occupancy | 1 – 1000 |
| $\tau$ | Time since an infected person entered the room | 0-1000 h |
| $V$ | Room volume | $10-10^4 \text{ m}^3$ |
| $A$ | Floor surface area | 5–5,000 $\text{m}^2$ |
| $H$ | Mean ceiling height, $V/A$ | 2–6 m |
| $Q_a$ | Ventilation outflow rate | $1-10^5 \text{ m}^3/\text{h}$ |
| $\lambda_a$ | Ventilation (outdoor air exchange) rate, $Q_a/V$ | $0.1-30 \text{ h}^{-1}$ |
| $\lambda_r$ | Recirculation air exchange rate, $Q_r/V$ | $0.1-30 \text{ h}^{-1}$ |
| $p_f$ | Probability of droplet filtration via recirculation | 0–1.0 |
| $\lambda_f$ | Filtration removal rate, $p_f \lambda_r$ | $0-30 \text{ h}^{-1}$ |
| $p_m$ | mask penetration probability, $\bar{p}_m = p_m(\bar{r})$ | 0.01–0.1 |
| **Physical Parameters** |  |  |
| $r$ | Respiratory drop radius | $0.1-100 \mu\text{m}$ |
| $V_d$ | Drop volume, $\approx \frac{4}{3}\pi r^3$ | $10^{-5}-10^6 \mu\text{m}^3$ |
| $n_d$ | Drop number density per radius | $0.01-1.0 (\text{cm}^3 \mu\text{m})^{-1}$ |
| $v_s$ | Drop settling speed | $10^{-5}-10^2 \text{ mm/s}$ |
| $\lambda_s$ | Drop settling rate, $v_s(r)/H$ | $10^{-5}-10^2 \text{ h}^{-1}$ |
| $Q_b$ | Mean breathing flow rate | $0.5-3.0 \text{ m}^3/\text{h}$ |
| $C_0$ | Background $\text{CO}_2$ concentration | 250–450 ppm |
| $C_2$ | Exhaled $\text{CO}_2$ concentration | 0–40,000 ppm |
| $P_2$ | Production rate of exhaled $\text{CO}_2$ | $0.02-10 \text{ m}^3/\text{h}$ |
| **Epidemiological Parameters** |  |  |
| $S, I$ | Number of susceptible and infected persons | |
| $\beta_a$ | Airborne transmission rate per infected-susceptible pair | $10^{-6}-10 \text{ quanta/h}^{-1}$ |
| $\lambda_v$ | Pathogen (virion) deactivation rate | $0.01-10 \text{ h}^{-1}$ |
| $\lambda_c$ | Pathogen concentration relaxation rate, $\bar{\lambda}_c = \lambda_c(\bar{r})$ | $0.1-100 \text{ h}^{-1}$ |
| $\bar{r}$ | Effective infectious drop radius | $0.3-5 \mu\text{m}$ |
| $P$ | Pathogen production rate / air volume / drop radius | $10^{-6}-10^9 (\text{h}\mu\text{m})^{-1}$ |
| $C$ | Infectious pathogen concentration / air volume / radius | $10^{-8}-10^4 (\text{m}^3 \mu\text{m})^{-1}$ |
| $c_v$ | Pathogen (virion) concentration per drop volume | $10^4-10^{11} \text{ RNA copies/mL}$ |
| $c_i$ | Pathogen infectivity, $1/(\text{infectious dose})$ | 0.001–1.0 |
| $C_q$ | Infectiousness of breath, exhaled quanta concentration | $1-1000 \text{ quanta/m}^3$ |
| $\lambda_q$ | Quanta emission rate, $Q_b C_q$ | $1-1000 \text{ quanta/h}$ |
| $p_i$ | Probability a person is infected (prevalence) | 0–1 |
| $p_{im}$ | Probability a person is immune (by vaccination or exposure) | 0–1 |
| $p_s$ | Probability a person is susceptible, $p_s = 1 - p_i - p_{im}$ | 0–1 |
| $s_r$ | Relative susceptibility (or transmissibility) | 0.1–10 |
| $N_t$ | Expected number of infected-susceptible pairs | 0–1000 |
| $T_a$ | Expected number of airborne transmissions, $N_t \langle \beta_a \rangle \tau$ | 0–100 |
| $\mathcal{R}_{in}$ | Indoor reproductive number, $(N-1) \langle \beta_a \rangle \tau$ | 0.001–100 |
| $\epsilon$ | Risk tolerance, bound on $T_a$ | 0.005–0.5 |

The only epidemiological parameter in the guideline, *C*_*q*_, is the infectiousness of exhaled air, measured in units of infection quanta per volume for a given aerosolized pathogen. The notion of “infection quantum” introduced by Wells (1955) is widely used in epidemiology to measure the expected rate of disease transmission, which may be seen as a transfer of infection quanta between pairs of infected and susceptible individuals. For airborne transmission, a suitable concentration of infection quanta per volume, *C*_*q*_, can thus be associated with exhaled air without reference to the microscopic pathogen concentration. Notably, *C*_*q*_ is known to depend on the type of respiratory and vocal activity (resting, exercising, speaking, singing, etc.), being larger for the more vigorous activities (Buonanno et al., 2020b; Bazant and Bush, 2021) The relative susceptibility *s*_*r*_ is introduced as a scaling factor for *C*_*q*_ that accounts for differences in the transmissibility of different respiratory pathogens, such as bacteria or viruses (Rudnick and Milton, 2003; Li et al., 2008) with different strains (Volz et al., 2021; Davies et al., 2020), and for differences in the susceptibility of different populations, such as children and adults (Riediker and Morawska, 2020; Zhang et al., 2020a; Zhu et al., 2020).

The mask penetration probability, *p*_*m*_ *(r*), is a function of drop size that is bounded below by 0 (for the ideal limit of perfect mask filtration) and above by 1 (appropriate when no mask is worn). Standard surgical masks at low flow rates allow only 0.04-1.5% of the most infectious (submicron) aerosols to penetrate (Chen and Willeke, 1992), values that should be increased by a factor of 2 − 10 to account for imperfect fit (Oberg and Brosseau, 2008). Cloth masks show much greater variability (Konda et al., 2020). The mask penetration probability may also depend on respiratory activity (Asadi et al., 2020b) and direction of airflow (Pan et al., 2020). Here, for the sake of simplicity, we treat it as being constant over the limited aerosol size range of interest, and evaluate 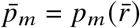 at the effective aerosol radius 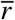 to be defined below. Bazant and Bush (2021) thus suggested 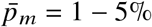 for surgical masks (Li et al., 2008; Oberg and Brosseau, 2008), 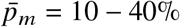 for hybrid cloth face coverings and 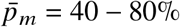 for single-layer fabrics (Konda et al., 2020). Notably, even low quality masks can significantly reduce transmission risk since the bound on cumulative exposure time, Eq. (1), scales as 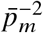.

Finally, we define 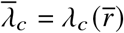 as an effective relaxation rate of the infectious aerosol-borne pathogen concentration, *C (r, t*), evaluated at the effective aerosol radius 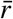. The size-dependent relaxation rate of the droplet-borne pathogen has four distinct contributions,

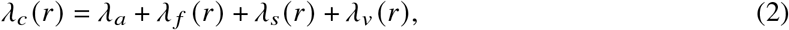

where *λ*_*a*_ is the ventilation rate (outdoor air exchanges per time). and *λ* _*f*_ *(r)* = *p*_*f*_ (r) *λ*_*r*_ is the filtration rate, where *p*_*f*_ *(r)* is the droplet removal efficiency for air filtration at a rate *λ*_*r*_ (recirculated air changes per time). *λ*_*s*_ *(r*) = *v*_*s*_ *(r) A*/*V* is the net sedimentation rate for infectious droplets with the Stokes settling velocity *v*_*s*_ *(r)* sedimenting through a well-mixed ambient to a floor of area A (Corner and Pendlebury, 1951; Martin and Nokes, 1988). Finally, *λ*_*v*_ *(r)* is the deactivation rate of the aerosolized pathogen, which depends weakly on humidity and droplet size (Yang and Marr, 2011; Lin and Marr, 2019; Marr et al., 2019), and may be enhanced by other factors such as ultraviolet (UV-C) irradiation (Hitchman, 2021; García de Abajo et al., 2020), chemical disinfectants (Schwartz et al., 2020), or cold plasma release (FilipiΔ et al., 2020; Lai et al., 2016).

Notably, only the first of the four removal rates apparent in Eq. (2) is relevant in the evolution of CO_2_; thus, the concentrations of CO_2_ and airborne pathogen are not strictly slaved to one another. Specifically, the proportionality between the two equilibrium concentrations varies in different indoor settings (Peng and Jimenez, 2021), for example in response to room filtration (Hartmann and Kriegel, 2020). Moreover, when transient effects arise, for example, following the arrival of an infectious individual or the opening of a window, the two concentrations adjust at different rates. Finally, we note that there may also be sources of CO_2_ other than human respiration, such as emissions from animals, stoves, furnaces, fireplaces, or carbonated beverages, as well as sinks of CO_2_, such as plants, construction materials or pools of water, which we neglect for simplicity. As such, following Rudnick and Milton (2003), we assume that the primary source of excess CO_2_ is exhalation by the human occupants of the indoor space.

In order to prevent the growth of an epidemic, the safety guideline should bound the indoor reproductive number, *ℛ*_*in*_, which is the expected number of transmissions if an infectious person enters a room full of susceptible persons. Indeed, the safety guideline, Eq. (1), corresponds to the bound *ℛ*_*in*_ < *ϵ* with the choice *N*_*t*_ = *N* − 1, and so would limit the risk of an infected person entering the room of occupancy *N* transmitting to any other during the exposure time *τ*. If the epidemic is well underway or subsiding, the guideline should take into account the prevalence of infection *p*_*i*_ and immunity *p*_*im*_ (as achieved by previous exposure or vaccination) in the local population. Assuming a trinomial distribution of *N* persons who are infected, immune or susceptible, with mutually exclusive probabilities *p*_*i*_, *p*_*im*_ and *p*_*s*_ = 1 − *p*_*i*_ *−p*_*im*_, respectively, the expected number of infected-susceptible pairs is *N (N* −1) *p*_*i*_*p*_*s*_. It is natural to switch between these two limits (*N*_*t*_ = *N* − 1 and N_*t*_ = *N (N* − 1) *p*_*i*_*p*_*s*_) when one infected person is expected to be in the room, *Np*_*i*_ = 1, and thus set

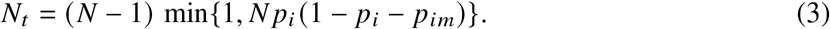

One may thus account for the changing infection prevalence *p*_*i*_ and increasing immunity *p*_*im*_ in the local population as the pandemic evolves.

### 2.2. CO_2_-based safety guideline

The total rate of CO_2_ production by respiration in the room is given by *p*_2_ = *NQ*_*b*_*C*_2,*b*_, where *C*_2,*b*_ is the CO_2_ concentration of exhaled air, approximately *C*_2,*b*_ = 38, 000 ppm, although the net CO_2_ production rate, *Q*_*b*_*C*_2,*b*_ varies considerably with body mass and physical activity (Persily and de Jonge, 2017). If the production rate *p*_2_ and the ventilation flow rate *Q* = *λ*_*a*_*V* are constant, then the steady-state value of the excess CO_2_ concentration, relative to the steady background concentration *C*_0_ at zero occupancy, is given by

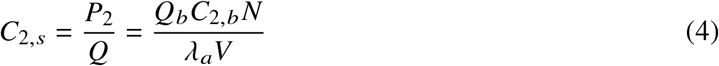

which is simply the ratio of the individual CO_2_ flow rate, *Q*_*b*_*C*_2,*b*_, to the ventilation flow rate per person, *Q*/*N*. We note that the outdoor CO_2_ concentration is typically in the range *C*_0_ = 250 − 450 ppm, with higher values in urban environments (Prill et al., 2000). In the absence of other indoor CO_2_ sources, human occupancy in poorly ventilated spaces can easily lead to CO_2_ levels of several thousand ppm. People have reported headaches, slight nausea, drowsiness, and decreased decision-making performance for levels above 1000 ppm (Fisk et al., 2013; Krawczyk et al., 2016), while short exposures to much higher levels may go unnoticed. As an example of CO_2_ limits in industry, the American Conference of Governmental Industrial Hygienists recommends a limit of 5000 ppm for an 8-hour period and 30,000 ppm for 10 minutes. A value of 40, 000 ppm is considered to be immediately life-threatening.

The safety guideline, Eq. (1), was derived on the basis of the conservative assumption that the infectious aerosol concentration has reached its maximum, steady-state value. If we assume, for consistency, that the CO_2_ concentration has done likewise, and so approached the value expressed in Eq. (4), then the guideline can be recast as a bound on the safe mean excess CO_2_ concentration,

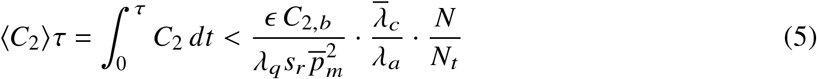

where we replace the steady excess CO_2_ concentration with its time average, ⟨*C*_2_ ⟩ ≈ *C*_2,*s*_, and define the mean quanta emission rate per infected person, *λ*_*q*_ = *Q*_*b*_*C*_*q*_. For the early to middle stages of an epidemic or when *p*_*i*_ and *p*_*im*_ are not known, we recommend setting *N*/*N*_*t*_ = 1 < *N*/ (*N* − 1) ≈ 1, for a conservative CO_2_ bound that limits the indoor reproductive number. In the later stages of an epidemic, as the population approaches herd immunity (*p*_*i*_ *→*0, *p*_*im*_ *→*1), the safe CO_2_ bound diverges, *N*/*N*_*t*_ *→ ∞*, and so may be supplanted by the limits on carbon dioxide toxicity noted above, that lie in the range 5, 000 − 30, 000 ppm for 8-hour and 10-minute exposures.

Our simple CO_2_-based safety guideline, Eq. (5), reveals scaling laws for exposure time, filtration, mask use, infection prevalence and immunity, factors that are not accounted for by directives that would simply impose a limit on CO_2_ concentration. The substantial increase in safe occupancy times, as one proceeds from the peak to the late stages of the pandemic, is evident in the difference between the solid and dashed lines in Figure 1, which were evaluated for the case of a typical classroom in the United States (Bazant and Bush, 2021). This example shows the critical role of exposure time in determining the safe CO_2_ level, a limit that can be increased dramatically by mask use and to a lesser extent by filtration. When infection prevalence p_*i*_ falls below 10 per 100,000 (an arbitrarily chosen small value), the chance of transmission is extremely low, allowing for long occupancy times. The risk of transmission at higher levels of prevalence, as may be deduced by interpolating between the solid and dashed lines in Figure 1, could also be rationally managed by monitoring the CO_2_ concentration and adhering to the guideline.

**Figure 1.**
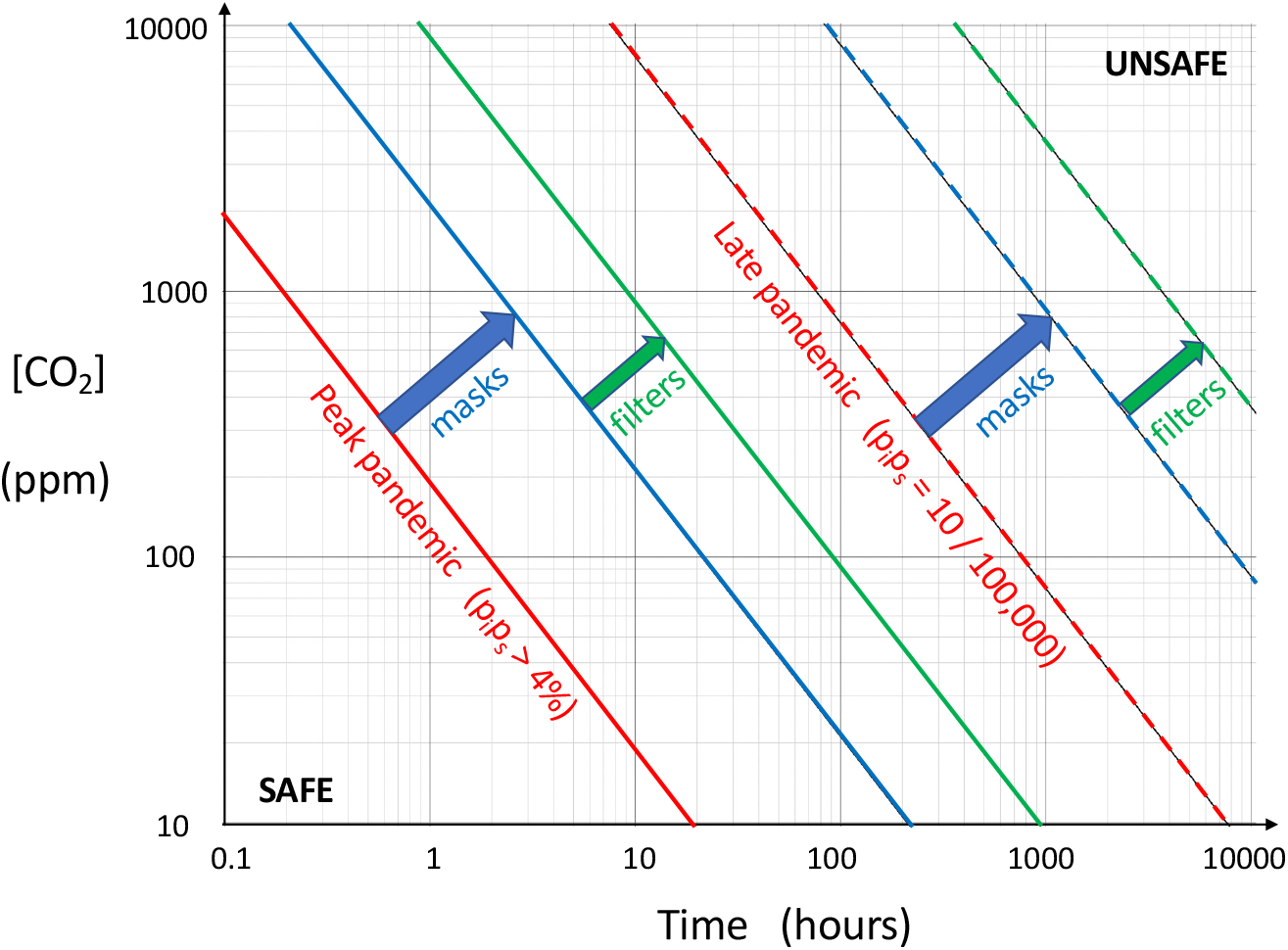
Illustration of the safety guideline, Eq. (5), which bounds the safe excess CO_2_ (ppm) and exposure time g (hours). Here, we consider the case of a classroom with N = 25 occupants, assumed to be children engaging in normal speech and light activity (λ_q_ s_r_ = 30 quanta/h) with moderate risk tolerance (*ϵ* = 10%). Compared to the most restrictive bound on the indoor reproductive number without any precautions (red line), the safe CO_2_ level or occupancy time is increased by at least an order of magnitude by the use of face masks (blue line), even with relatively inconsistent use of cloth masks (p_m_ = 30%). The effect of air filtration (green line) is relatively small, shown here for a case of efficient HEPA filtration (p_f_ = 99%) with 17% outdoor air fraction (λ_f_ = 5λ_a_). All three bounds are increased by several orders of magnitude (dashed lines) during late pandemic conditions (p_i_ p_s_ = 10 per 100, 000), when it becomes increasingly unlikely to find an infected-susceptible pair in the room. The other parameters satisfy (λ _ν_ + λ_s_)/ λ_a_ = 0.5, as could correspond to, for example, λ _ν_= 0.3/h, λ_B_ = 0.2/h and λ_a_ = 1/h (1 ACH).

## 3. Mathematical Model of CO_2_ Monitoring to Predict Airborne Disease Transmission Risk

### 3.1. CO_2_ dynamics

We follow the traditional approach of modeling gas dynamics in a well mixed room (Shair and Heitner, 1974), as a continuous stirred tank reactor (Davis and Davis, 2012). Given the time dependence of occupancy, *N*(*t*), mean breathing flow rate, *Q*_*b*_ (*t*), and ventilation flow rate, *Q*_*a*_ (*t*) = *λ*_*a*_ (*t*)*V*, one may express the evolution of the excess CO_2_ concentration *C*_2_(*t*) in a well-mixed room through

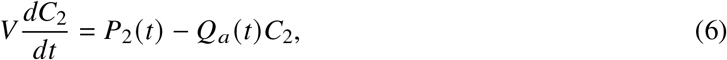

where

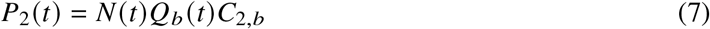

is the exhaled CO_2_ production rate. The relaxation rate for excess CO_2_ in response to changes in *P*_2_(*t*) is precisely equal to the ventilation rate, *λ*_*a*_ (*t*) = *Q*_*a*_ (*t*)/*V*. For constant *λ*_*a*_, the general solution of Eq. (6) for *C*_2_(0) = 0 is given by

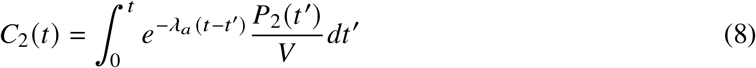

which can be derived by Laplace transform or using an integrating factor. The time-averaged excess CO_2_ concentration can be expressed as

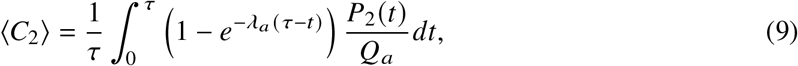

by switching the order of time integration. If *P*_2_ (*t*) is slowly varying over the ventilation time scale 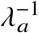, the time-averaged CO_2_ concentration may be approximated as

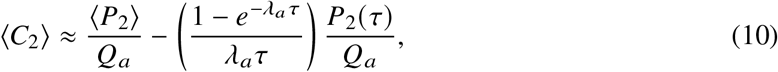

where the excess CO_2_ concentration approaches the ratio of the mean exhaled CO_2_ production rate to the ventilation flow rate at long times, 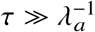, as indicated in Eq. (4).

### 3.2. Infectious aerosol dynamics

Following Bazant and Bush (2021), we assume that the radius-resolved concentration of infectious aerosol-borne pathogen, *C*(*r, t*), evolves according to

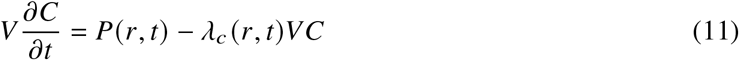

where the mean production rate,

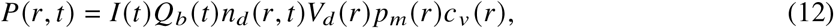

depends on the number of infected persons in the room, I (*t*), and the size distribution n_*d*_ (*r, t*) of exhaled droplets of volume *V*_*d*_ *(r)* containing pathogen (i.e. virions) at microscopic concentration, *c*_*ν*_ *(r)*. The droplet size distribution is known to depend on expiratory and vocal activity (Morawska et al., 2009; Asadi et al., 2019, 2020c). Quite generally, the aerosols evolve according to a dynamic sorting process (Bazant and Bush, 2021): the drop-size distribution evolves with time until an equilibrium distribution is obtained.

Given the time evolution of excess CO_2_ concentration, *C*_2_(*t*), one may deduce the radius-resolved pathogen concentration *C*(*r, t*) by integrating the coupled differential equation,

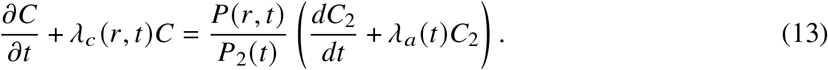

This integration can be done numerically or analytically via Laplace transform or integrating factors if one assumes that *λ*_*a*_, *λ*_*c*_, *P* and *P*_*2*_ all vary slowly over the ventilation (air change) time scale, 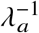. In that case, the general solution takes the form

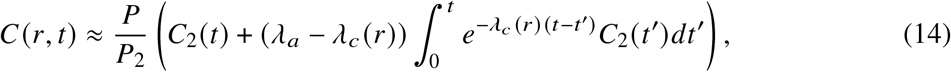

where we consider the infectious aerosol build-up from *C*(*r*, 0) = 0.

### 3.3. Disease transmission dynamics

According to Markov’s inequality, the probability of at least one transmission taking place during the exposure time *τ* is bounded above by the expected number of airborne transmissions, *T*_*a*_ (*τ*), and the two become equal in the (typical) limit of rare transmissions, *T*_*a*_ (*τ*) ≪ 1. The expected number of transmissions to *S*(*t*) susceptible persons is obtained by integrating the mask-filtered inhalation rate of infection quanta over both droplet radius and time,

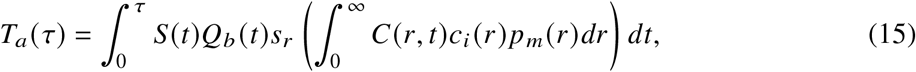

where *C*_*i*_ (r) is the infectivity of the aerosolized pathogen. The infectivity is measured in units of infection quanta per pathogen and generally depends on droplet size. One might expect pathogens contained in smaller aerosol droplets with *r* < 5 *μ*m to be more infectious than those in larger drops, as reported by Santarpia et al. (2020) for SARS-CoV-2, on the grounds that smaller drops more easily traverse the respiratory tract, absorb and coalesce onto exposed tissues, and allow pathogens to escape more quickly by diffusion to infect target cells. The mask penetration probability *P*_*m*_(*r*) also decreases rapidly with increasing drop size above the aerosol range for most filtration materials (Chen and Willeke, 1992; Oberg and Brosseau, 2008; Konda et al., 2020; Li et al., 2008), so the integration over radius in Eq. (15) gives the most weight to the aerosol size range, roughly *r* < 5*μ*m, which also coincides with the maxima in exhaled droplet size distributions (Morawska et al., 2009; Asadi et al., 2019, 2020c).

The inverse of the infectivity, 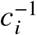, is equal to the “infectious dose” of pathogens from inhaled aerosol droplets that would cause infection with probability 1−(1/*e*) = 63%. Bazant and Bush (2021) estimated the infectious dose for SARS-CoV-2 to be on the order of ten aerosol-borne virions. Notably, the corresponding infectivity, *C*_*i*_ ∼0.1, is an order of magnitude larger than previous estimates for SARS-CoV (Watanabe et al., 2010; Buonanno et al., 2020b), which is consistent with only COVID-19 reaching pandemic status. The infectivity is known to vary across different age groups and pathogen strains, a variability that is captured by the relative susceptibility, *s*_*r*_. For example, Bazant and Bush (2021) suggest assigning *s*_*r*_ = 1 for the elderly (over 65 years old), *s*_*r*_ = 0.68 for adults (aged 15-64) and *s*_*r*_ = 0.23 for children (aged 0-14) for the original Wuhan strain of SARS-CoV-2, based on a study of transmission in quarantined households in China (Zhang et al., 2020a). The authors further suggested multiplying these values by 1.6 for the more infectious variant of concern of the lineage B.1.1.7 (VOC 202012/01), which recently emerged in the United Kingdom with a reproductive number that was 60% larger than that of the original strain (Volz et al., 2021; Davies et al., 2020).

### 3.4. Approximate Formula for the Airborne Transmission Risk from CO_2_ Measurements

Equations (14) and (15) provide an approximate solution to the full model that depends on the exhaled droplet size distribution,*n*_*d*_(*r,t*), and mean breathing rate, *Q*_*b*_ (*t*), of the population in the room. Since the droplet distributions *n*_*d*_(*r*) A have only been characterized in certain idealized experimental conditions (Morawska et al., 2009; Asadi et al., 2020a,c), it is useful to integrate over *r* to obtain a simpler model that can be directly calibrated for different modes of respiration using epidemiological data (Bazant and Bush 2021). Assuming *Q*_*b*_(*t*), *I*(*t*), *S*(*t*), *N*(*t*) and *n*_*d*_(*r,t*)vary slowly over the relaxation time,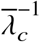 we may substitute Eq. (14) into Eq. (15) and perform the time integral of the second term to obtain

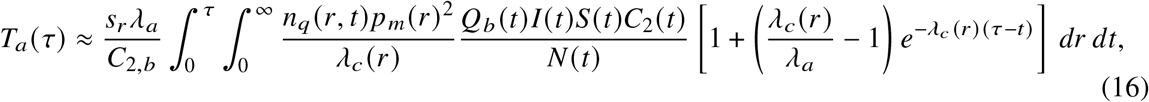

where *n*_*q*_(*r,t*)= *n*_*d*_(*r,t*)*V*_*d*_(*r*)*c*_*v*_(*r*)*c*_*i*_(*r*) is the radius-resolved exhaled quanta concentration.

Following Bazant and Bush (2021), we define an effective radius of infectious aerosols 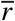 such that

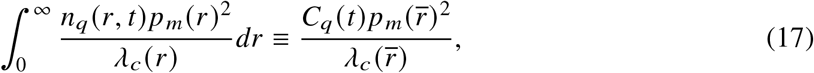

where 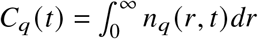 is the exhaled quanta concentration, which may vary in time with changes in expiratory activity, for example, following a transition from nose breathing to speaking. In principle, the effective radius 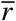 can be evaluated, given a complete knowledge of the dependence on drop radius of the mask penetration probability, *P*_*m*_(*r*), and of all the factors that determine the exhaled quanta concentration, *n*_*q*_(*r,t*)and pathogen removal rate, *λ*_*c*_(*r*). While these dependencies are not readily characterized, typical values of 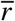 are at the scale of several microns, based on the size dependencies of *n*_*d*_(*r,t*), *c*_*i*_(*r*),and *p*_*m*_(*r*) noted above.

Further simplifications allow us to derive a formula relating CO_2_ measurements to transmission risk. By assuming that *C*_*q*_(*t*) varies slowly over the timescale of concentration relaxation, one may approximate the memory integral with the same effective radius 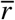. Thus, accounting for immunity and infection prevalence in the population via

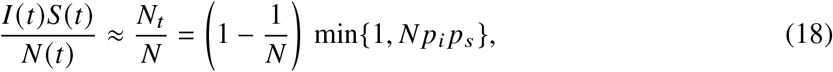

we obtain a formula for the expected number of airborne transmissions,

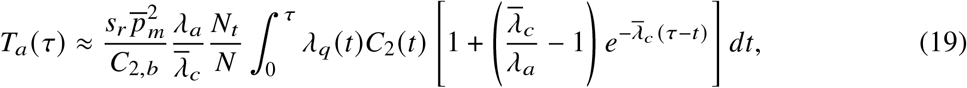

in terms of the excess CO_2_ time series, *C*_2_(*t*), where *λ*_*q*_(*t*) = *C*_*q*_(*t*)*Q*_*b*_(*t*)is the mean quanta emission rate. It is also useful to define the expected transmission rate,

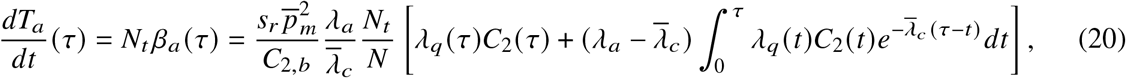

which allows for direct assessment of airborne transmission risk based on CO_2_ levels. A pair of examples of such assessments will be presented in §4.

Notably, the mean airborne transmission rate per expected infected-susceptible pair, *β*_*a*_(*t*), reflects the environment’s memory of the recent past, which persists over the pathogen relaxation time scale,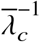 The CO_2_ concentration in Eq. (9) has a longer memory of past changes in CO_2_ sources or ventilation, which persists over the air change time scale, 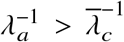 since CO_2_ is unaffected by the filtration, sedimentation and deactivation rates evident in Eq. (2). The time delays between the production of CO_2_ and infectious aerosols by exhalation and their buildup in the well mixed air of a room shows that CO_2_ variation and airborne transmission are inherently non-Markovian stochastic processes. As such, any attempt to predict fluctuations in airborne transmission risk would require stochastic generalizations of the differential equations governing the mean variables, Eqs. (6) and (11), and so represent a stochastic formulation of the Wells-Riley model (Noakes and Sleigh, 2009).

### 3.5. Reduction to the CO_2_-based Safety Guideline

Finally, we connect the general result, Eq. (19), with the CO_2_ based safety guideline derived above, Eq. (5). Since *C*_2_(t) varies on the ventilation time scale, 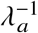, which is necessarily longer than the relaxation time scale of the infectious aerosols, 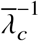, we may assume that *λq*(*t*)_2_*C*_2_(*t*) is slowly varying and evaluate the integral in Eq. (19). We thus arrive at the approximation

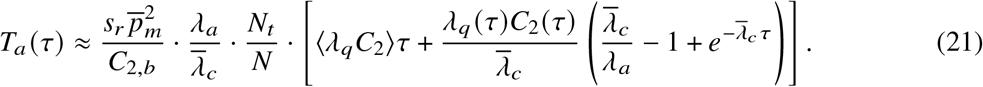

Since *λ*_*q*_(*t*) *C*_2_(*t*) is slowly varying, the second term in brackets is negligible relative to the first for times longer than the ventilation time, 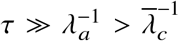. In this limit, the imposed bound on expected transmissions, *T*_*a*_ (*τ*) < *ϵ*, is approximated by

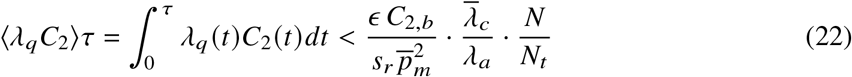

This formula reduces to the safety guideline, Eq. (5), in the limit of constant mean quanta emission rate, *λ*_*q*_, which confirms the consistency of our assumptions.

## 4. Examples

We proceed by illustrating the process by which the guideline, Eq. (5), can be coupled to real data obtained from CO_2_ monitors. Specifically, we consider time series of CO_2_ concentration gathered in classroom and in office settings at the Massachusetts Institute of Technology using an Atlas Scientific EZO-CO2 Embedded NDIR CO2 Sensor controlled with an Arduino Uno, and an Aranet4, respectively. Social distancing guidelines were adhered to, and masks were worn by all participants. We assume a constant exhaled CO_2_ concentration of 38, 000 ppm, and use the global minimum of the CO_2_ series as the background CO_2_ level *C*_0_ from which the excess concentration *C*_2_ (*t*) was deduced. Notably, the relatively small fluctuations in the CO_2_ measurements recorded in a variety of settings support the notion of a well-mixed room.

From Equations (19) and (20), we calculate the expected number of transmissions and transmission rate, assuming that there is one infected person in the room (*N*_*t*_ = *N* − 1). In this case, the expected number of transmissions is equal to the indoor reproductive number, T_*a*_ (*τ*) = ℛ_*in*_(*τ*), and the transmission rate is 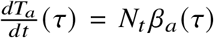. The approximations in the derivation are valid in these examples, so a direct numerical solution of Eq. (13) would yield indistinguishable results. In particular, the “slowly varying” assumptions are satisfied, since we keep *N* and *I* constant, and any time-dependence of *Q*_*b*_ cancels in the ratio *P* /*P*_2_. The droplet distributions *nd* (*r,t*) A, C may vary in time, but no significant changes in mean respiratory activity were observed or measured. Moreover, accounting for any variations in *λ*_*a*_ and *λ*_*c*_ would require additional measurements in the same space, so these parameters may also be taken as constants during the relatively short exposure times considered.

We choose realistic values of the parameters that fall within the typical ranges estimated by Bazant and Bush (2021). The mean breathing rate is set to *Q*_*b*_ = 0.5 *m*^3^ / *h* for light activity, and 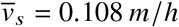 for sedimentation with 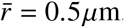. The viral deactivation rate for SARS-CoV-2 is set to *λ*_*v*_ = 0.3 / *h*, an estimate appropriate for 50% relative humidity. We plot *N*_*t*_ *β*_*a*_ (*τ*) and *ℛ*_*in=*_ (*τ*) for cases where masks are and are not worn, and choose a mask penetration probability of *p*_*m*_ ≈ 0.3, as is roughly appropriate for a cloth mask. The parameters 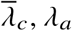, *H* and *N* are chosen according to the specific scenario presented. Exhaled COVID-19 quanta concentrations *Cq* for various expiratory activities are estimated from Fig. 2 of Bazant and Bush (2021). In order to be conservative, we assume that s_*r*_ = 1, suitable for the high-risk individuals exposed to the Wuhan strain of SARS-CoV-2.

**Figure 2.**
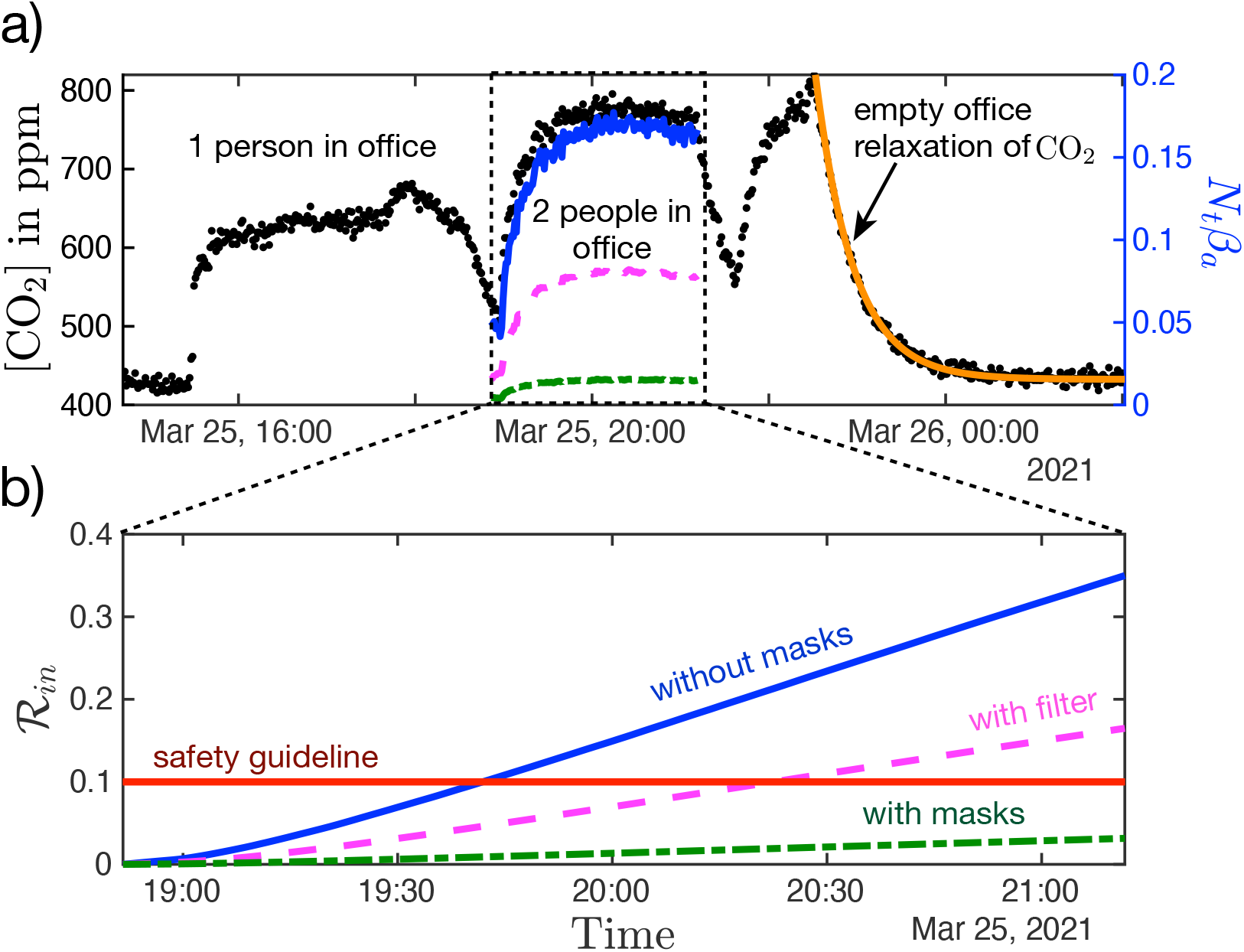
Measured CO_2_ concentration and calculated transmission rate in a two-person office. a) Black dots represent the concentration of CO_2_. The solid blue, dashed magenta and dash-dot green curves represents the transmission rate, as calculated from (20) for three different scenarios, two of which were hypothetical: (blue) the pair are not wearing masks and there is no filtration present; (magenta) the pair are not wearing masks and there is filtration present; (green) the pair are wearing masks and there is no filtration present. The orange solid curve denotes the period of exponential relaxation following the exit of the room’s occupants, from which one may infer the room’s ventilation rate, λ_a_ = 2.3 /h. b) Corresponding blue, magenta and green curves, deduced by integrating (20), indicate the total risk of transmission over the time of shared occupancy. If the pair were not wearing masks, the safety limit ℛ_in_ < 0.1 would be violated after approximately an hour.

**Figure 3.**
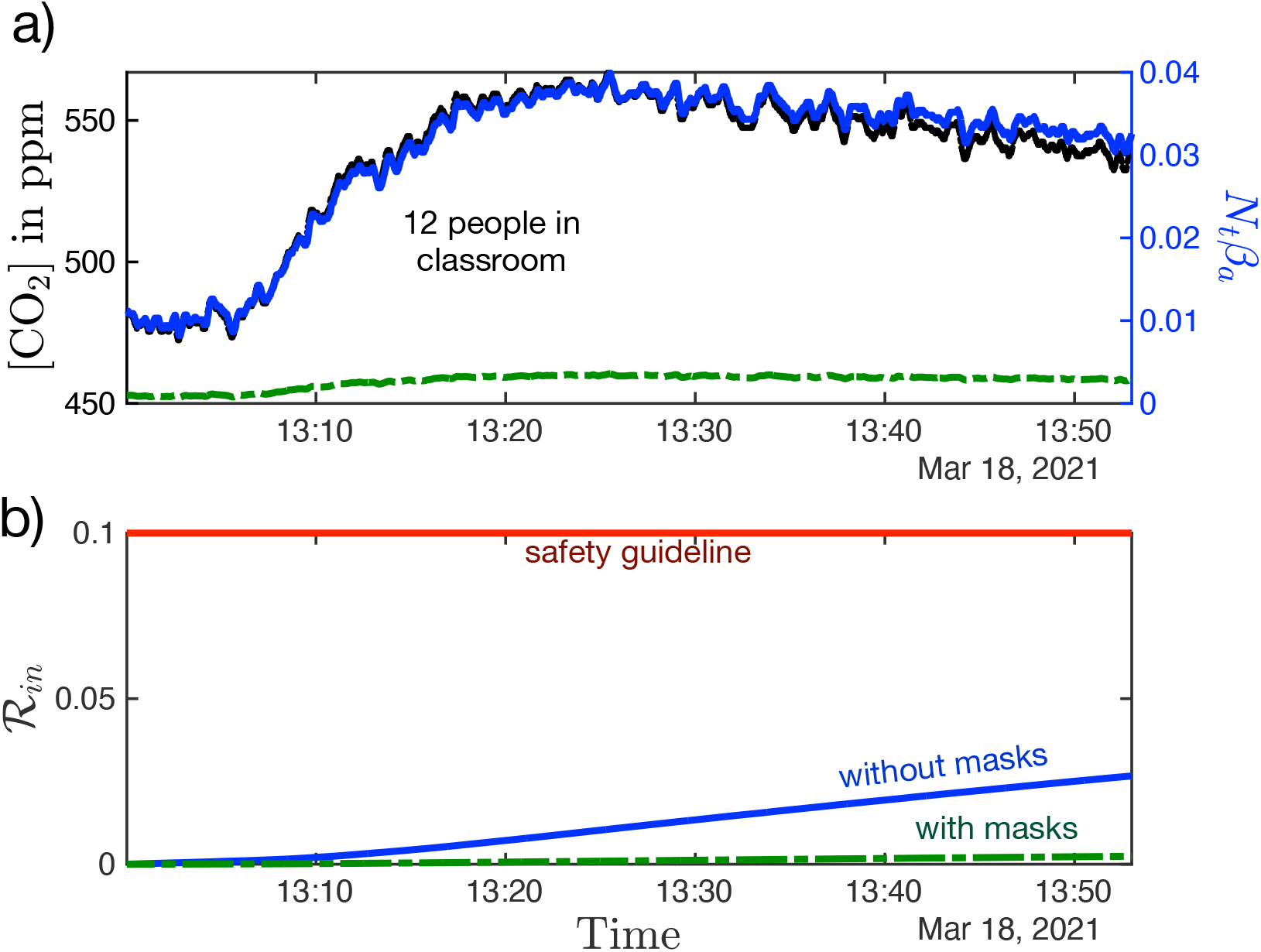
Measured CO_2_ concentration and calculated transmission rate for 12 masked students in a university lecture hall. a) Black dots represent the concentration of CO_2_. The solid blue and dash-dot green curves represent the transmission rate, as calculated from (20), when the occupants are wearing masks, and in the hypothetical case where they are not, respectively. b) The solid blue and dash-dot green curves indicate the total risk of transmission with and without masks, respectively, as deduced by integrating (20) over time. Even had masks not been worn, the safety guideline would not have been violated during the lecture.

### 4.1. Small office with two workers

Figure 2(a) shows CO_2_ measurements taken in an office of length *L* = 4.2 m, width, *W*= 3 m, and height *H* = 3 m. Initially, a single worker is present, but at 19:00, a second worker arrives at the office. The workers exit the office at 21:09, return at 21:39, and exit again at 22:30. As the participants were speaking, we use *C*_*q*_= 72 quanta /*m*^3^. We compute the room’s ventilation rate, *λ*_*a*_ 2.3 / *h*, from the exponential relaxation that follows the occupants’ exit from the office (as indicated by the orange curve in Figure 2). The office was equipped with moderate ventilation, which we characterize with *p*_*f*_ = 0.99 and *λ*_*r*_ = 6 / *h*.

As shown in Figure 2(b), if the office mates were not wearing masks, the safety guideline of expected transmissions *T*_*a*_ < 10% would be violated after approximately an hour together. However, office mates wearing cloth masks remain well under the safety guideline during the 2.5 hours spent together. Filtration without masks also extends the occupancy time limits, however ℛ_*in*_ approaches the safety limit after approximately 2 hours. While this example should not be taken as a definitive statement of danger or safety in this setting, it does serves to illustrate how our CO_2_ guideline can be implemented in a real-life situation.

### 4.2. University classroom adhering to social distancing guidelines

We next monitor CO_2_ levels during a university lecture. There were *N*= 12 participants in a lecture hall of length *L* = 13 m, width, *W*= 12 m and height *H* = 3 m. The lecture started at 13:05 and finished at 13:50. Four people remained in the room for 30 minutes after class. The classroom has mechanical ventilation, which we characterize in terms of *λ*_*a*_ = 2 / *h* (30 minute outdoor air change). During the lecture, the professor spoke while the students were quiet and sedentary; thus, we assume *C*_*q*_ = 30 quanta /*m*^3^. The maximum CO_2_ concentration reached in the classroom was ≈550 ppm, the excess level no more than 100 ppm. Thus, the safety limit was never exceeded, and would not have been even if masks had not been worn. We note that this lecture hall was particularly large, well-ventilated and sparsely populated, and so should not be taken as being representative of a classroom setting.

A more complete assessment of safety in schools would require integrating CO_2_ concentrations over a considerably longer time interval. For example, if students are tested weekly for COVID-19, then one should assess the mean CO_2_ concentration in class during the course of an entire school week. Nevertheless, this second example further illustrates the manner in which our model may be applied to real-world settings and suggests that precautions such as ventilation, filtration and mask use, can substantially increase safe occupancy times.

## 5. Conclusion

Mounting evidence suggests that COVID-19 is spread primarily via indoor airborne transmission. Such an inference is no surprise, as such is also the case for many other respiratory illnesses, including influenza, tuberculosis, measles and severe-acute respiratory syndrome (as is caused by a precursor to SARS-CoV-2, the coronavirus SARS-CoV). More than a year into the pandemic, public health guidance continues to emphasize the importance of social distancing and surface cleaning, despite evidence that mask directives are much more effective than either in limiting airborne transmission. We have here illustrated the manner in which CO_2_ monitoring may be used in conjunction with the safety guideline of Bazant and Bush (2021) in assessing the risk of indoor airborne respiratory disease transmission, including that of COVID-19. We thus hope to inform personal and policy decisions about closing and re-opening indoor spaces, such as schools and businesses.

We have here reformulated the COVID-19 indoor safety guideline of Bazant and Bush (2021), expressing it in terms of cumulative exposure to carbon dioxide, which can be readily monitored in real time for most indoor spaces. In so doing, we have built upon the important early work of Rudnick and Milton (2003), as was recently extended and applied to COVID-19 by Peng and Jimenez (2021). The guideline of Bazant and Bush (2021) makes clear that, since the risk of indoor airborne infection is determined by the total volume of pathogen inhaled, safety limits intended to protect against it must be expressed in terms of occupancy time. Likewise, in the context of CO_2_ measurements, safety limits cannot be expressed solely in terms of a limit on carbon dioxide levels, but must also depend on occupancy time. As we have demonstrated with our two case studies, the safety guideline (5), when coupled with CO_2_ monitors, allows for real-time assessment of risk of airborne disease transmission ℛ_*in*_ in indoor spaces. Moreover, this approach has the distinct advantage that one can assess certain key model parameters, including the background concentration of CO_2_ and the room’s ventilation rate, directly from the CO_2_ measurements.

Within a well-mixed space, carbon dioxide is effectively a passive scalar that tracks the ambient flow, and is removed only through exchange with outdoor air. Aerosol-borne pathogen is subject to additional removal mechanisms, including filtration (by face masks and internal circulation), sedimentation and deactivation. Thus, the concentration of CO_2_ cannot be taken as a proxy for that of aerosol-borne pathogen without resolving the proportionality constant between the two that results from these additional removal mechanisms. We stress that the effects of face mask use are dramatic in reducing the ratio of aerosol-borne pathogen to CO_2_ concentration, and so in reducing the risk of indoor transmission. Finally, we note that the additional removal mechanisms acting on the droplet-borne pathogen alter not only its equilibrium concentrations relative to that of CO_2_, but their relaxation times in transient situations, as may be treated using the mathematical formalism of Bazant and Bush (2021).

We emphasize that the caveats enumerated by Bazant and Bush (2021) concerning the limitations of their safety guideline apply similarly here. First, there is considerable uncertainty in a number of model parameters, including the critical viral load and relative susceptibility. While our inferences are consistent with available data, we hope that these uncertainties will be reduced as more COVID-19 spreading events are characterized and analyzed. Second, when masks are not worn, there is substantial additional risk of short-range airborne transmission from respiratory jets and plumes, as accompany breathing, speaking (Abkarian et al., 2020a; Abkarian and Stone, 2020), coughing and sneezing (Bourouiba et al., 2014). Third, while the assumption of the well-mixed room is widely applied and represents a reasonable first approximation, it is known to have limitations (Bhagat et al., 2020; Linden et al., 1990; Linden, 1999). Notably, measured fluctuations in CO_2_ levels provide a direct means of assessing the validity of the well-mixed-room hypothesis, especially when several sensors are used simultaneously at different locations in the same space. Indeed, CO_2_ monitoring has been used for decades to assess the quality of air handling and zonal mixing in buildings (Seppänen et al., 1999; Fisk and De Almeida, 1998; Hung and Derossis, 1989; Salisbury, 1986; Cheng et al., 2011), and can now be re-purposed to assess the risk of indoor airborne disease transmission.

Our transmission theory and safety guideline provide a quantitative basis for the use of CO_2_ monitors in assessing the risk of indoor airborne disease transmission. Specifically, the simple guideline, Eq. (5), and mathematical formulae connecting CO_2_ data to the evolving transmission risk, Eqs. (19)-(20), pave the way for real-time assessment of personal risk in indoor spaces. Moreover, our results should allow for building-scale optimization of public health using CO_2_ sensors, wherein risk assessment might be weighed against the energy requirements of enhanced ventilation. Finally, our model provides a general framework for using CO_2_ monitors to mitigate the indoor airborne transmission of other respiratory illnesses, including the seasonal flu.

Extensions of our study would include implementing our guideline in spaces where not only CO_2_ is monitored, but also the room occupancy along with other relevant parameters appearing in our model. For example, monitoring decibel levels and type of vocalization could serve to inform the infectious aerosol production rate (Asadi et al., 2019, 2020c; Morawska et al., 2009). Changes in occupancy could also be monitored at entrances and exits (while maintaining anonymity). One could further envision feeding all such data into air regulation controls in order to ensure that our CO_2_-based indoor safety limit is never violated. We note that such a prospect would be most easily achieved in quasi-steady circumstances in which a fixed population is behaving in a predictable fashion over the course of an event of known duration, for example, for students in a lecture hall or passengers on a charter bus. In more complicated situations, our model provides a framework for optimizing sensor-based demand-controlled ventilation (Fisk and De Almeida, 1998) with a view to limiting transmission risk while reducing energy consumption and system costs.

In order to facilitate the application of our safety guideline, in the Supplementary Material we provide a link to an online app that computes the safety guideline in terms of both room occupancy and CO_2_ levels (Khan et al., 2020). The CO_2_-based guideline, available in the app’s Advanced Mode, may be used in conjunction with CO_2_ monitors to formulate safe re-opening policies for indoor spaces in the later stages of the pandemic.

## Data Availability

The data presented in the article is available upon request.

https://indoor-covid-safety.herokuapp.com

## Acknowledgements

We thank Laura Champion for valuable discussions and for sharing preliminary data.

## Author Contributions

J.W.M.B. and M.Z.B. conceived the study. M.Z.B. developed the mathematical model. O.K. performed the experiments and analyzed the data with A.E.C. K.K. developed the online app with M.Z.B and J.W.M.B. All authors contributed to writing the manuscript.

## Data Availability Statement

Raw data are available from M.Z.B. and O.K.

## Ethical Standards

The research meets all ethical guidelines, including adherence to the legal requirements of the study country.

## Supplementary Material

An online app is provided to compute the COVID-19 Indoor Safety Guideline, where the safe CO_2_ level versus time can be displayed in Advanced Mode (Khan et al., 2020).

